# Anaemia Types and Severity in Patients aged 1 to 14 years at the Children’s Hospital of the University Teaching Hospitals in Zambia

**DOI:** 10.1101/2020.09.17.20194779

**Authors:** Panji Nkhoma, Patrick Loti, Musalula Sinkala, Hamakwa Mantina, Florence Mwaba, Doris Kafita, Oliver Mwenechanya, Sody Munsaka

## Abstract

Anaemia is a condition in which either the number of red blood cells or their oxygen-carrying capacity is insufficient to meet physiologic needs, which vary by age, sex, altitude, smoking and pregnancy status. The global estimate of childhood anaemia indicates that 293.1 million children are anaemic, and 28.5% of these children reside in sub-Sahara Africa. Also, anaemia is a significant public health problem with a high age-standardised death rate of 11.18 per 100,000 in Zambia.

We conducted a cross-sectional study involving 392 anaemic children aged one year to 14 years. The study was conducted at the Children Hospital, University Teaching Hospitals, which is a third-level referral Hospital in Lusaka, Zambia. The aim was to determine the most common type of anaemia, it’s severity, and the most affected age groups among children aged 1–14 years.

Out of 392 participants, 219 (56%) were female. Maximum haemoglobin recorded was 10.9g/dl, a minimum of 2.0 g/dl, a mean of 7.8g/dl and a standard deviation of 1.86g/dl. 200 (51%) participants had severe anaemia, and 192 (49%) had moderate anaemia with none having mild anaemia. Microcytic hypochromic anaemia was the commonest (60%), followed by normochromic normocytic anaemia (26%) and the least was macrocytic anaemia in 14% of the participants. An analysis of variance (ANOVA) showed that the difference in mean haemoglobin concentration between age groups was not significant, F (7.94) = 0.83, p > 0.57. A Chi-squared test was used to determine the relationship between anaemia types (microcytic, hypochromic) and age groups. The interaction was not significant (Chi-Square (1) = 1.28, p-value = 0.73. Microcytic hypochromic anaemia was the most prevalent and all age groups were equally affected. We recommend the country’s National Food and Nutrition Commission to revisit the Zambian National Strategy and Plan of Action for the Prevention and Control of Vitamin A Deficiency and Anaemia of 1999 to 2004 and implement the measures stated in the strategic plan.

## INTRODUCTION

Anaemia is a condition in which the number of red blood cells or their oxygen-carrying capacity is insufficient to meet physiologic needs, which vary by age, sex, altitude and pregnancy status [1]. Clinically, physicians consider anaemia whenever there are reductions in haematocrit and/or haemoglobin, the latter of which is a surrogate value for anaemia [2]. The World Health Organization (WHO) has set reference cut-off points for healthy populations and defines anaemia as a public health problem if more than 20% of the population is anaemic, whereas the prevalence of greater than 40% is considered as a serious public health problem [3]. The global estimate of childhood anaemia indicates that 293.1 millions of children are anaemic, and 28.5% of these children reside in sub-Sahara Africa [4] which Zambia is part of. Also, anaemia is regarded as a significant public health problem in Africa because it is the cause of death for most children admitted to hospitals [5]. Anaemia can occur at all stages of human life, although young children and pregnant women are the most affected, with an estimated global prevalence of 43% and 51% respectively [6].

Anaemia can be caused by a multitude of factors [7]. These factors may be genetic, such as haemoglobinopathies; infections, such as malaria, intestinal helminths and chronic infection; or nutritional, which includes iron deficiency as well as deficiencies of other vitamins and minerals, such as folate, vitamins A and B12 and copper [8]. The two leading causes are deficiency in micronutrients and parasitic infections. Iron deficiency is the main micronutrient that contributes to anaemia [9,10]. Anaemia in children is of particular significance and interest since if untreated may lead to mental impairment, physical and social development, all of which culminate to adverse behavioural and cognitive effects that impact the child’s performance in school and/or their work capacity in adulthood [11]. The study aimed to determine the most common type of anaemia, it’s severity, and the most affected age groups among children aged 1-14 years admitted in Children’s Hospital at University Teaching Hospitals, Lusaka, Zambia.

## METHODOLOGY

### Study design

The study was cross-sectional involving anaemic children aged one year to 14 years at the Children Hospital, University Teaching Hospitals, which is a third-level referral and largest hospital in Zambia.

### Experimental Approach

Blood was collected in Ethylenediaminetetraacetic acid (EDTA) specimen container for a full blood count. The samples were run within thirty minutes to an hour after collection using Sysmex XT 4000i (Sysmex Corporation, Germany) haematology analyser. All quality control procedures were run to ensure the reliability of results. Classification of anaemia was made using red blood cell indices (MCV and MCH values)

### Data Analysis

Data were entered in Microsoft Excel and analysed using python 3.7 for Mac. Age characteristics, type of anaemia and severity of anaemia were analysed using descriptive statistics and presented as means, frequencies, percentages and graphs. Data were checked for normality using the Shapiro Wilk Test and log-transformed so that it is near normal distribution. Analysis of Variance (ANOVA) was used to determine whether there were mean haemoglobin differences among age groups and the Chi-squared test was used to determine the relationship between types of anaemia, severity and age groups. P-values of < 0.05 were considered statistically significant.

### Ethical Considerations

The study was approved by the University of Zambia Health Sciences Research Ethics Committee (UNZAHSREC); Protocol ID No: 20190217064.

## RESULTS

### Demographic Characteristics

We recruited 392 participants out of which 219 (56%) were female. The age of the participants ranged from 1 year to 14 years with a mean age of 6.1 years and a standard deviation of 4.2 years. The median age of the participants was 5 years. The majority of the participants were toddlers (42%) aged 1 to 3 years old, preschoolers(16%) aged 4 to 6 years old, school-aged (29%) aged 7 to 11 years old and adolescents(13%) aged 12 to 14 years (Figure 1).

**Figure 1:**
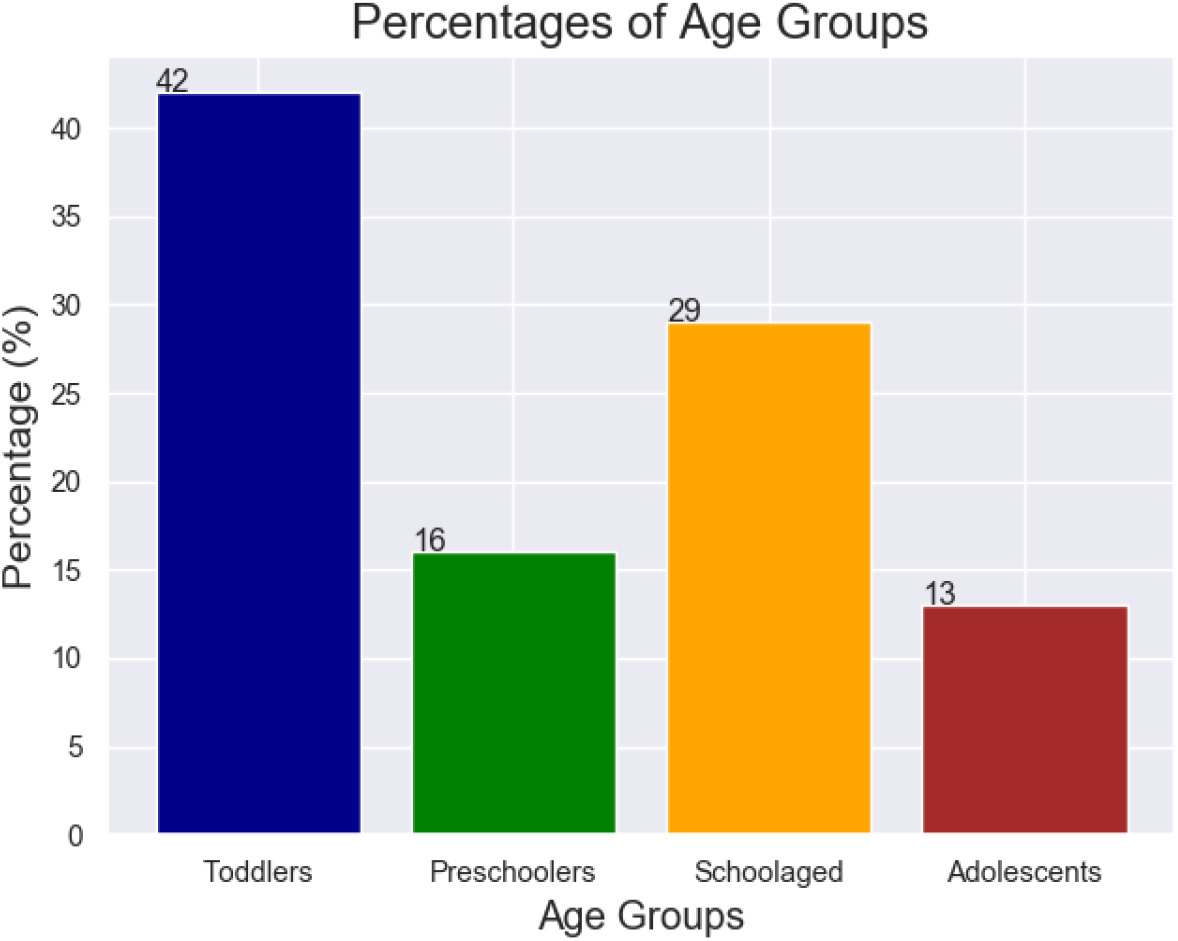
Showing distribution of participants by age groups. Toddlers were the most followed by school-aged children

### Anaemia Types and Severity

The highest haemoglobin level that we recorded was 10.9 g/dl, whereas the minimum was 2.0 g/dl. We found a mean haemoglobin level of 7.8 g/dl, a median of 8 g/dl and a standard deviation of 1.86 g/dl. Here also, we showed that 200 (accounting for 51%) of participants had severe anaemia, and 192 (49%) had moderate anaemia.

Microcytic hypochromic anaemia was the most prevalent type of anaemia 234 (60%), followed by normochromic normocytic anaemia 103 (26%) and then macrocytic anaemia in 55 (14%) of the participants.

### Mean Haemoglobin Differences Between Age Groups

We categorised the study participants in four groups as follows: (1) toddlers (1 to 3 years old), (2) preschooler (4 to 6 years old), (3) school-aged (6 to 11 years old) and (4) adolescents (above 12 years old). Then we compared differences in the mean haemoglobin concentration using a one-way Analysis of variance (ANOVA). Interestingly, we found no statistically significant difference in the mean haemoglobin concentration between our four categories of participants, F (7.94) = 0.83, p > 0.57 (Figure 2).

**Figure 2:**
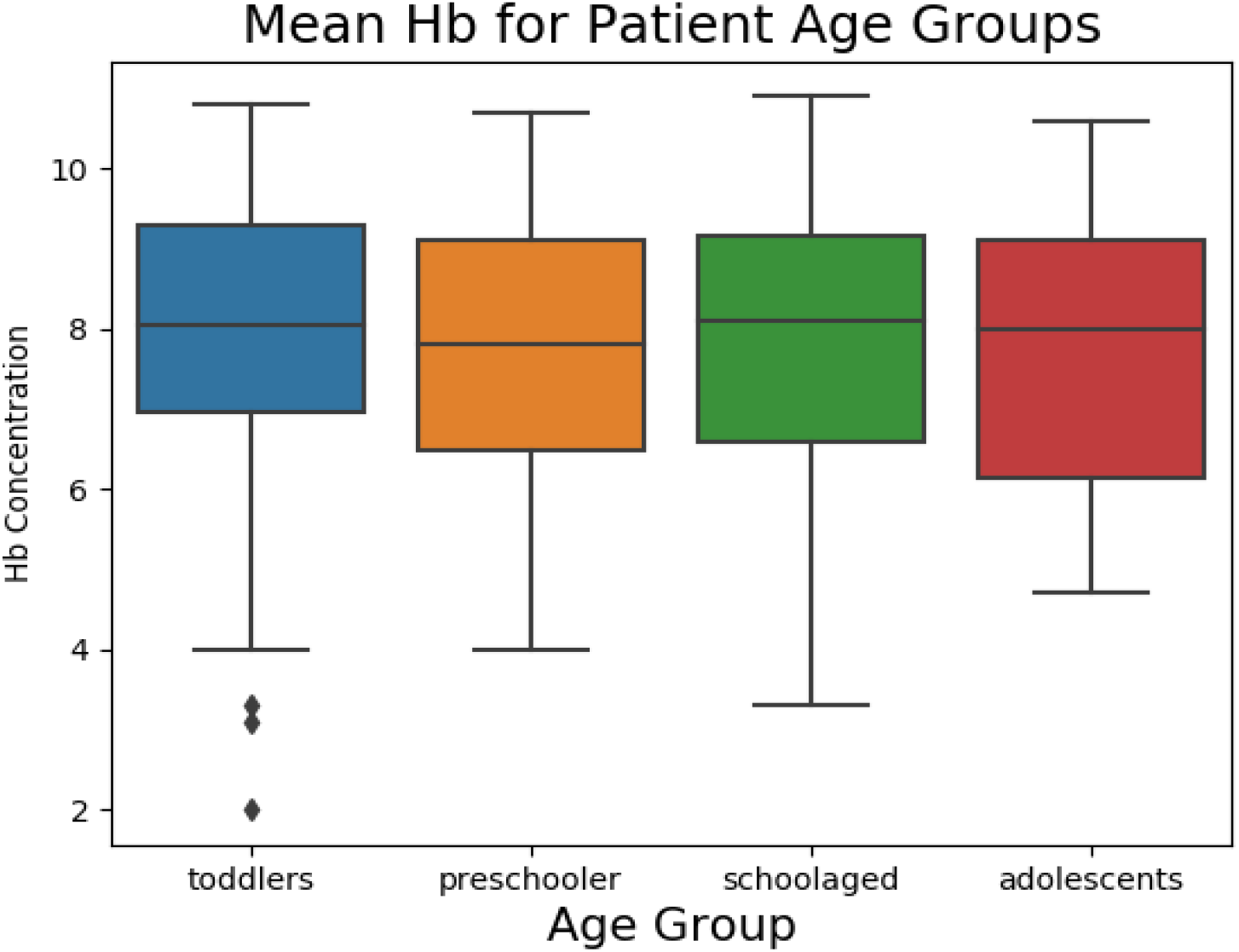
Showing Mean Haemoglobin Differences between Age Groups. The mean differences were not statistically different

### Relationship Between Anaemia Types, severity and Age Groups

We performed a Chi-squared test of independence to determine the relationship between anaemia type, severity and the categories of participants age groups that we have earlier defined. Here, we found no significant interaction Chi-Square = 1.28, p-value = 0.73 and Chi-square = 1.27, p-value = 0.74 respectively.

## DISCUSSION

Generally, anaemia has, for a long time, been a problem in Zambian children [12]. However, the country has recorded a steady decrease in the prevalence of anaemia in children under 5 years old from about 75% in 1990 to about 54% in 2016 [12]. Despite the reduction, the prevalence of 54% is still of severe public health significance according to WHO guidelines [13]. We have revealed that microcytic-hypochromic anaemia, which is mainly caused by iron deficiency in the affected individuals, is the most prevalent type of anaemia affecting our study participants of all age groups. This is not surprising as microcytic-hypochromic anaemia is reported to be the most common form of anaemia globally [14], and in most of these cases, iron deficiency is the primary cause of anaemia [9,15–17]. The deficiency of iron in children usually comes about due to inadequate iron supply to the body as a consequence of, among others, low dietary iron, and/or low bioavailability of dietary iron, increased iron requirements due to growth, and increased iron losses due to gastrointestinal blood loss and diarrhoea [18–20]. Notably, in the early 2000s, iron deficiency was reported to be the most common micronutrient deficiency in Zambia, which affected millions of people, including school children and adolescents [21]. Now, almost 20 years on, this study still shows high iron deficiency among the study participants. This problem persists despite the many strategies aimed to alleviate iron deficiency such as iron fortification of staple food and iron supplementation pills [21]. Since iron is required for optimal growth and development [22], iron deficiency can lead to impaired neural development and motor and cognitive function, increased risk of mortality and reduced productivity [23]. Therefore, if the problem is not looked into from a new perspective and solved, this will likely adversely affect the intellectual capacity of children and the future workforce of Zambia and many other sub-Saharan countries that are affected.

Here, we report that all children that we examined had moderate to severe anaemia. Since anaemia is recognised as the major cause of death among children either directly [24] or as a contributing risk factor [25], our findings suggest that this is a problem that should be looked at with serious concern.

This distribution of moderate to severe anaemia in our study could be because the University Teaching Hospital where the study was conducted is a level 3 referral hospital receiving patients whose conditions cannot be handled by lower levels of hospitals in the city. Furthermore, Zambia is a developing country where most households may not afford foods rich in iron and other minerals required for the formation of blood cells [26]. This study was conducted in an urban area and agreed with findings of some studies that found that children in such areas are more likely to be anaemic [27]. Surprisingly, all age groups defined in our study are affected equally in terms of anaemia type and severity. This shows that no age group is spared as all are probably facing similar problems that lead to anaemia.

We recommend the country’s National Food and Nutrition Commission to revisit the Zambian National Strategy and Plan of Action for the Prevention and Control of Vitamin A Deficiency and Anaemia of 1999 to 2004 and implement the measures stated in the strategic plan.

## CONCLUSION

Microcytic hypochromic anaemia was the most prevalent with no significant mean difference in haemoglobin levels across all ages. All age groups ranging from toddlers to teens were affected equally in terms of anaemia types and severity. Also, anaemia ranged from moderate to severe.

## DATA AVAILABILITY

The data used to support the findings of this study are available from the corresponding author upon request

## AUTHOR CONTRIBUTIONS

Conceptualisation: Panji Nkhoma, Musalula Sinkala, Sody Munsaka, Patrick Loti, Hamakwa Mantina, Doris Kafita

Data Analysis: Panji Nkhoma, Musalula Sinkala, Sody Munsaka, Doris Kafita.

Investigation: Panji Nkhoma, Musalula Sinkala, Sody Munsaka, Patrick Loti, Florence Mwaba, Doris Kafita, Oliver Mwenechanya, Hamakwa Mantina

Supervision: Panji Nkhoma, Musalula Sinkala, Sody Munsaka, Florence Mwaba, Hamakwa Mantina

Writing original draft: Panji Nkhoma, Doris Kafita, Florence Mwaba, Sody Munsaka, Musalula Sinkala, Hamakwa Mantina

